# Practical alcohol risk-reduction advice plus a brief commitment declaration in a social drinking laboratory: a pilot cluster randomized trial

**DOI:** 10.64898/2026.04.19.26351067

**Authors:** Hisashi Yoshimoto, Takahiro Hadano, Kensuke Shimada, Masahiko Gosho, Takafumi Fukuda, Yuta Komano, Kentaro Umeda, Masamori Iwase, Yuko Kusano, Takayo Kawabata

**Author notes:** Corresponding author: Hisashi Yoshimoto, Research and Development Center for Lifestyle Innovation, University of Tsukuba, 1-2 Kasuga, Tsukuba, Ibaraki 305-8550, Japan, Department of Family Medicine, General Practice and Community Health, Institute of Medicine, University of Tsukuba, 1-1-1 Tennodai, Tsukuba, Ibaraki 305-8575, Japan.

## Abstract

**Background:** Practical alcohol risk-reduction strategies are widely recommended in public-facing alcohol guidance, but randomized evidence from socially interactive drinking episodes remains limited. We conducted a pilot cluster randomized trial to evaluate the feasibility and preliminary effects of a package intervention comprising practical drinking-strategy information, participant self-selection of same-day strategies, and a brief commitment declaration in a social drinking laboratory.

**Methods:** This single-center, parallel-group pilot trial was conducted in Japan. Pre-existing social groups participated. One or two groups scheduled in the same session slot were combined into a time-slot allocation unit, which was randomized 1:1 either to the package intervention or to alcohol-related knowledge only. The primary outcome was total pure alcohol intake during the first 120 min. Session satisfaction on a Visual Analog Scale (VAS) was a prespecified secondary participant-experience outcome.

**Results:** Of 83 interested individuals, 63 were randomized and 59 participants in 17 social groups and 12 allocation units were included in the modified intention-to-treat analysis. The mean paired intervention-control difference for 120-min alcohol intake was-8.84 g (95% confidence interval [CI]-27.92 to 10.23; exact sign-flip p = 0.281). The corresponding exploratory 0–30 min difference was-4.90 g (95% CI-10.48 to 0.68; exact sign-flip p = 0.094). In a genotype-adjusted participant-level sensitivity analysis, the intervention coefficient for 120-min intake was-16.0 g (95% CI-30.9 to-1.1; p = 0.036). Session satisfaction was high in both arms with no clear between-arm difference. Next-day follow-up was 100%, and no adverse-event-related discontinuations occurred.

**Conclusions:** The intervention was feasible to deliver in a socially interactive drinking setting, and session satisfaction was high in both arms. Primary allocation-unit estimates favored lower alcohol intake but were imprecise. Larger trials are needed to estimate effects more precisely, while considering the potential influence of genotype imbalance on effect estimation in East Asian samples.

**Trial registration:** University Hospital Medical Information Network Clinical Trials Registry (UMIN-CTR) UMIN000060685. Registered 17 February 2026.

## 1. Background

Alcohol-related harm remains a major public health concern, and the risks associated with drinking depend not only on how much is consumed but also on the context in which drinking occurs. Event-level reviews indicate that drinking quantity is shaped by social, environmental, and person-level factors acting together, suggesting that laboratory paradigms that approximate real drinking situations may help evaluate practical public-health advice in behaviorally relevant settings [1,2].

Human laboratory alcohol studies have traditionally provided tightly controlled tests of subjective intoxication, craving, reward, and self-administration, but many paradigms have been based on solitary drinking or minimally social conditions [3]. More recent theoretical and review work emphasizes that alcohol reward is strongly influenced by social context and that social drinking situations may be critical for understanding how behavior changes during a drinking episode [4,5].

One relevant conceptual framework for such advice is protective behavioral strategies (PBS), particularly “manner of drinking” strategies such as drinking more slowly, spacing drinks, and alternating alcoholic with non-alcoholic beverages. Reviews indicate that PBS are generally associated with less heavy drinking and fewer alcohol-related consequences [6,7]. However, the evidence base is still dominated by observational studies. Randomized studies of PBS-related interventions have shown mixed findings: some multicomponent interventions that include PBS have reduced weekly or peak drinking, whereas standalone PBS-focused interventions have not consistently produced clear direct reductions in alcohol use or consequences [8–11].

The gap between widely disseminated practical advice and direct experimental evidence is also evident in national drinking guidelines. Public-facing guidance in Japan, the United Kingdom, Australia, and Canada commonly recommends practical strategies such as deciding a limit in advance, drinking more slowly, eating while drinking, alternating alcoholic and non-alcoholic drinks, and choosing lower-alcohol beverages [12–15]. However, direct experimental evidence on the immediate effects of individual strategies within social drinking episodes remains limited [8–11]. Thus, practical advice is widely disseminated even though the behavioral mechanisms and immediate effects remain incompletely tested.

The current study therefore examined whether a package intervention combining practical drinking-strategy information, participant self-selection of same-day strategies, and an on-site commitment declaration could change alcohol intake in a social drinking laboratory setting. This pilot aimed to estimate feasibility and preliminary effect sizes for a brief approach to communicating practical alcohol risk-reduction advice. Because alcohol metabolism and flushing are strongly influenced by alcohol dehydrogenase 1B (ADH1B) and aldehyde dehydrogenase 2 (ALDH2) in East Asian populations [16], we also examined in sensitivity analyses whether accounting for these variants materially altered the intervention effect estimate. We hypothesized that the intervention would reduce total alcohol intake and, because several strategies target early pacing of drinking, we also explored whether any signal might emerge in the early part of the drinking episode.

## 2. Methods

### 2.1 Study design and setting

This was a single-center, parallel-group, pilot cluster randomized trial conducted in a social drinking laboratory in Japan. Participants enrolled as pre-existing self-formed social groups rather than as individually recruited strangers. These self-formed social groups were the units for participation and for individual-level mixed-effects analyses, whereas allocation units were defined as time-slot-based randomization units that could include one or two self-formed groups. To minimize contamination, one or two self-formed groups scheduled within the same time slot were co-assigned to the same study condition; thus, the time-slot allocation unit was the operational unit of randomization. Enrollment and study sessions were conducted from February 18 to 23, 2026, with next-day follow-up completed by February 24, 2026. As a pilot study, sample size was determined pragmatically by recruitment and scheduling feasibility rather than by a formal power calculation.

### 2.2 Participants

Eligible participants were adults aged 20 years or older who had previously consumed more than 100 g of pure alcohol in a day without major health problems, had consumed at least 60 g of pure alcohol on one occasion within the past month, could participate with at least two other people, were classified as non-flushers on the flushing questionnaire [17], and were able to provide written informed consent. Participants who were pregnant or breastfeeding, intoxicated before the session, unable to provide informed consent, or judged to have alcohol dependence were excluded. For participants with Alcohol Use Disorders Identification Test (AUDIT) scores ≥15 [18], a physician experienced in alcohol-dependence care conducted an interview and determined eligibility based on International Classification of Diseases, 11th Revision (ICD-11) criteria.

### 2.3 Cluster formation and randomization

Self-formed social groups were intended to include 3–5 participants; groups of six or more were split as evenly as possible. For randomization, one or two self-formed groups scheduled in the same drinking-session time slot were combined into a single time-slot allocation unit and assigned together to the same study condition. Each allocation unit therefore represented one scheduled drinking-session slot and could contain one or two self-formed social groups. Allocation units, not self-formed groups, were the units of randomization. Usual-drinking values used for block construction were obtained from a web questionnaire completed after eligibility screening and before schedule finalization and randomization. After schedules were finalized, the 12 allocation units were ranked using the mean usual-drinking value within each allocation unit, and adjacent units were paired into 6 randomization blocks; each block contained two allocation units matched on mean usual drinking. The random allocation sequence was generated using Research Randomizer, and within each paired block one allocation unit was assigned to the intervention and the other to the control condition. Participant eligibility and scheduling were implemented by study staff according to predefined web-screening and scheduling criteria, and allocation of the time-slot units to study condition was implemented according to the generated sequence after schedule finalization.

### 2.4 Masking, intervention procedures, and safety monitoring

Participants were informed of a general study theme, but the specific hypothesis was not disclosed until all study procedures were complete. Before drinking began, participants completed baseline measurements, including age, sex, height, weight, and recent alcohol use, and then received a condition-specific information session lasting approximately 5–10 min. Day-of-session measurement staff were not given the allocation schedule in advance, and participants were not shown the allocation schedule. Because the intervention involved condition-specific materials and a commitment declaration, this was an open-label behavioral comparison and masking could not be maintained throughout the drinking session. The primary outcome, however, was based on staff-recorded beverage residual volumes rather than subjective ratings.

The intervention was a package intervention comprising practical drinking-strategy information, participant self-selection of one or more strategies to use that day, and a required commitment declaration. The control condition provided alcohol-related knowledge only. No additional behavioral co-intervention was delivered after the condition-specific information session.

Participants who withdrew before study procedures therefore did so before any condition-specific materials were delivered and before they were informed of their assigned condition. The practical strategies were deciding the amount to drink that day in advance (with an optional planned pure-alcohol target entry), drinking slowly during the first 30 min, eating while drinking, alternating alcoholic drinks with water or non-alcoholic drinks, choosing lower-alcohol beverages, and putting the drink down on the table between sips. During the 120-min ad libitum drinking period, participants could order beverages through study staff, and beverages were not self-served. Food items such as pizza, fries, chicken, and salad were placed on the table before drinking began and were not separately ordered. Canned beverages were served in their original containers, red wine was kept at room temperature, and white wine was chilled and served by staff in 100-mL pours. Alcoholic beverages were collected every 30 min for residual-volume checks used to derive participant-level alcohol consumption; alcohol-free beverages and soft drinks were collected and measured at 120 and 180 min. Beverage sharing was not permitted.

Sessions were monitored continuously by at least one physician or medically trained staff member. Prespecified stopping criteria included vomiting, marked physical discomfort, disruptive intoxicated behavior, physician judgment that continuation was unsafe, or participant request. To reduce fall risk, staff accompanied participants to the restroom as needed.

Participants remained under observation during the recovery period and returned home by taxi under predefined departure procedures.

### 2.5 Outcomes and genotyping

The primary outcome for the present manuscript was total pure alcohol intake over 120 min. A prespecified secondary participant-experience outcome was session satisfaction on a Visual Analog Scale (VAS) assessed at 120 min. Pure alcohol intake was derived for each participant from the volume of each beverage consumed during each 30-min interval and converted to grams of pure alcohol using beverage-specific alcohol concentration and volume. Trained study staff checked beverage remnants at each 30-min interval and recorded residual volumes for the alcoholic beverages. Usual alcohol consumption was assessed using a categorical questionnaire based on the traditional Japanese unit gou (1 gou ≈ 20 g of ethanol) [12]. Participants selected a category of usual intake, and these categories were converted to estimated grams/day using category midpoints (e.g., 10 g/day for <1 gou and 30 g/day for 1–<2 gou, with 20-g increments thereafter). Saliva samples were collected at baseline for ADH1B and ALDH2 genotyping.

### 2.6 Statistical analysis

The primary analysis set was a modified intention-to-treat (mITT) sample including randomized participants who started study procedures and provided primary outcome data. No imputation was performed. Participants who withdrew before study procedures and provided no outcome data were excluded from the mITT set; among participants who started study procedures, primary outcome data were complete. Statistical analyses were conducted independently by an academic researcher who was not employed by the sponsor and had no sponsor-related financial conflicts of interest, using Stata/MP 19.0 and R 4.5.2. The study plan and analysis approach were reviewed in advance by an academic biostatistician. In genotype-adjusted models, ADH1B and ALDH2 were entered as categorical variables, with *1/*1 as the reference category for each gene. The primary inferential analyses were allocation-unit analyses aligned with the unit of randomization [19]. Allocation-unit means were calculated for each block, intervention–control differences were derived within blocks, and these paired block differences were summarized with t-based 95% confidence intervals (CIs) and exact sign-flip tests [20]. Because allocation-unit sizes varied, we also report block-size-weighted sensitivity analyses, weighting each block-specific intervention–control difference (D_b) by the total number of analyzed participants in that block (N_b); the corresponding exact sign-flip sensitivity statistic was T = Σ(N_b × D_b). This sensitivity analysis gives larger blocks more influence, but it does not estimate an exact participant-average treatment effect when arm sizes differ within blocks. The primary estimand for allocation-unit analyses remained the unweighted allocation-unit average effect, because the allocation unit was the unit of randomization and we did not want larger units to contribute more weight by default [21]. Individual-level mixed-effects models with random intercepts for self-formed groups were conducted as sensitivity analyses. Session satisfaction VAS was analyzed as a prespecified secondary participant-experience outcome using allocation-unit analyses for between-arm comparison and participant-level mixed models as sensitivity analyses. Exploratory genotype interaction analyses were also fitted. Because strategy selections within the intervention arm were self-selected, overlapping, and highly uneven, all strategy selections were first summarized descriptively in the supplementary materials. To avoid overinterpreting sparse or highly uneven selections, focused exploratory models were then limited to early-pacing-related selections with non-sparse uptake and direct conceptual relevance to early-interval intake. Early interval intake was a prespecified conceptual interest of the intervention, but interval-specific statistical testing was treated as exploratory. No formal progression criteria were prespecified.

### 2.7 Ethics and registration

The study was approved by the Institutional Ethics Committee for Human Genome and Genetic Research, University of Tsukuba (approval numbers G316 and G316-1) and prospectively registered on February 17, 2026, with the University Hospital Medical Information Network Clinical Trials Registry (UMIN-CTR; UMIN000060685). Summaries of intervention materials, safety procedures, and protocol-to-analysis deviations are provided in the supplementary materials. During manuscript preparation, the authors used ChatGPT (OpenAI) to translate draft text from Japanese into English, assist with English-language editing, and obtain mock reviewer-style feedback on manuscript clarity and presentation. All outputs were reviewed and edited by the authors, who take full responsibility for the content of the manuscript.

## 3. Results

### 3.1 Participant flow and feasibility

Among 83 individuals who expressed interest, 20 were excluded before randomization (7 because groups did not meet the minimum size requirement and 13 because scheduling capacity was exceeded). These 20 individuals belonged to 9 self-formed groups. This left 63 participants in 18 self-formed groups. At randomization, 31 participants in 9 self-formed groups were assigned to the intervention arm and 32 participants in 9 self-formed groups were assigned to the control arm. After randomization but before study procedures, one control-arm self-formed group (3 participants) withdrew because of family circumstances and one additional control-arm participant withdrew because of influenza infection. Thus, 59 participants in 17 self-formed groups were included in the modified intention-to-treat analysis. The analyzed self-formed groups ranged from 3 to 5 participants in size (mean 3.5). The next-day follow-up response rate was 100%, and no participant discontinued because of adverse events or physician-initiated stopping. Although one control-arm self-formed group withdrew before study procedures, its time-slot allocation unit still contained another participating self-formed group; therefore, all 12 allocation units remained analyzable. The four participants who withdrew before study procedures had not yet been informed of their assigned condition.

### 3.2 Baseline characteristics and genotype distributions

Baseline characteristics are shown in Table 1. Mean usual drinking was 69.3 g/day in the control arm and 55.2 g/day in the intervention arm. ADH1B genotype distribution did not differ between arms (Fisher’s exact test, p = 0.295). ALDH2 genotype distribution differed between arms (p = 0.008), with ALDH2*1/*2 observed in 35.7% of the control arm and 6.5% of the intervention arm. These p values are reported only to document the observed genotype distribution for interpretation of genotype-adjusted sensitivity analyses [22].

**Table 1.**
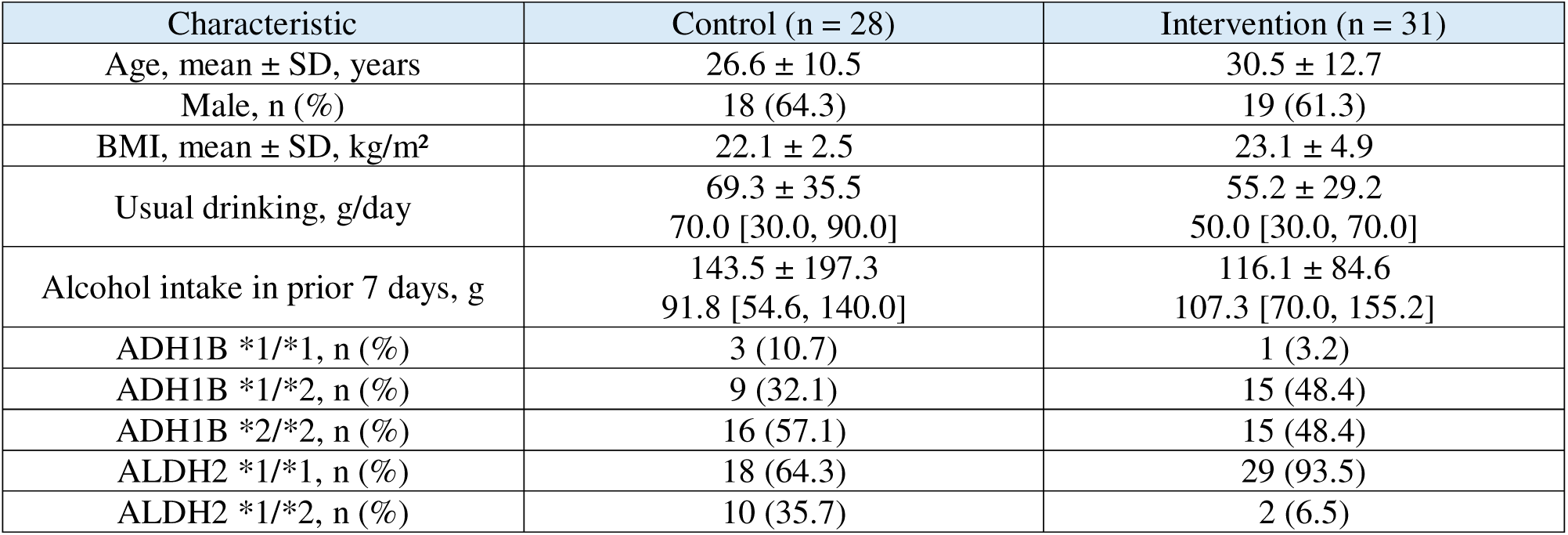
Baseline characteristics and genotype distributions in the modified intention-to-treat sample.

### 3.3 Allocation-unit analysis of the primary outcome

The allocation-unit analysis included 12 allocation units (6 intervention and 6 control) across 6 randomization blocks. Allocation units contained a mean of 4.9 participants (range 3–8) and comprised one or two self-formed groups. For the primary outcome of 120-min total pure alcohol intake, the mean paired intervention–control difference across blocks was-8.84 g (95% CI-27.92 to 10.23; exact sign-flip p = 0.281). The corresponding block-size-weighted summary difference, weighting each block-specific difference by the total number of analyzed participants in that block, was-8.95 g, and the block-size-weighted exact sign-flip sensitivity analysis yielded the same p value (0.281). For exploratory 0–30 min intake, the mean paired intervention–control difference was-4.90 g (95% CI-10.48 to 0.68; exact sign-flip p = 0.094). The corresponding block-size-weighted summary difference was-5.02 g, and the block-size-weighted exact sign-flip p value was 0.125. Block-level allocation-unit means, allocation-unit composition, and sensitivity results are summarized in Supplementary Table S2.

### 3.4 Individual-level mixed-effects sensitivity analyses

For descriptive participant-level comparison, mean 120-min pure alcohol intake was 56.6 g in the intervention arm and 66.8 g in the control arm. Individual-level mixed-effects models with random intercepts for self-formed groups were fitted as sensitivity analyses (Table 2). In the unadjusted model, the intervention coefficient was-7.3 g (95% CI-28.4 to 13.8, p = 0.498).

**Table 2.** Primary allocation-unit results and individual-level mixed-model sensitivity analyses.

| Panel | Analysis/model | Covariates included in model | Intervention estimate (g) | 95% CI | p value | N |
| --- | --- | --- | --- | --- | --- | --- |
| **Panel A** | Allocation-unit analysis: 120-min total intake | None; paired block differences in allocation-unit means | −8.84 | −27.92 to 10.23 | 0.281* | 6 blocks |
|  | Allocation-unit exploratory analysis: 0–30 min intake | None; paired block differences in allocation-unit means | −4.90 | −10.48 to 0.68 | 0.094* | 6 blocks |
| **Panel B** | Model 1 | None | −7.29 | −28.36 to 13.78 | 0.498 | 59 |
|  | Model 2 | Usual drinking | −2.24 | −21.33 to 16.85 | 0.818 | 59 |
|  | Model 3 | Usual drinking, age, sex, BMI | −5.86 | −22.15 to 10.44 | 0.481 | 59 |
|  | Model 4 | Usual drinking, age, sex, BMI, ADH1B, ALDH2 | −15.99 | −30.91 to −1.08 | 0.036 | 59 |

\*Exact sign-flip p value. Panel A shows allocation-unit analyses aligned with the unit of randomization; estimates are paired block differences in allocation-unit means (intervention – control). For Panel A, 95% CIs are t-based intervals for block-difference means. Block-size-weighted exact sign-flip sensitivity p values were 0.281 for 120-min total intake and 0.125 for exploratory 0–30 min intake; corresponding block-level allocation-unit values are given in Supplementary Table S2. Panel B shows participant-level mixed-effects models with random intercepts for self-formed groups; estimates are intervention coefficients from the fitted models.

After adjustment for usual drinking (Model 2), the coefficient was-2.2 g (95% CI-21.3 to 16.9, p = 0.818). After further adjustment for age, sex, and body mass index (BMI) (Model 3), the coefficient was-5.9 g (95% CI-22.2 to 10.4, p = 0.481). In a sensitivity analysis additionally adjusting for ADH1B and ALDH2 (Model 4), the coefficient was-16.0 g (95% CI-30.9 to-1.1, p = 0.036). Exploratory genotype interaction analyses are summarized in Supplementary Table S3 as interaction coefficients relative to the reference genotype categories. The intraclass correlation coefficient (ICC) for 120-min total intake at the self-formed-group level in the unadjusted mixed-effects model was 0.33 (Supplementary Table S3).

### 3.5 Exploratory interval-specific analyses, session satisfaction, and descriptive strategy uptake summaries

Exploratory Model 4 interval-specific mixed-model results are shown in Supplementary Figure S2, and participant-level mean ± standard deviation (SD) values by study arm are shown in Supplementary Figure S3. Session satisfaction VAS, a prespecified secondary outcome, was high in both arms (mean 87.8 in the intervention arm and 91.7 in the control arm). In allocation-unit analyses, the mean paired intervention–control difference was-3.98 points (95% CI-18.77 to 10.82; exact sign-flip p = 0.531). In a participant-level mixed-effects sensitivity analysis with random intercepts for self-formed groups, the intervention coefficient was-3.33 points (95% CI - 11.50 to 4.83; p = 0.424). Across all six self-selected strategies in the intervention arm, uptake was heterogeneous, with no uptake for the advance limit-setting strategy and near-universal uptake for eating while drinking; descriptive frequencies and crude contrasts for 0–30 min and 120-min total intake are shown in Supplementary Table S4 and Supplementary Figure S1.

Focused exploratory results for early-pacing-related selections are summarized in Supplementary Table S6. In within-intervention-arm models adjusted for usual drinking and alcohol intake during the prior 7 days, selecting “drink slowly during the first 30 min” was associated with lower alcohol intake during the first 30 min of the session (coefficient-5.88 g; 95% CI-10.07 to-1.69; p = 0.008). A similar pattern was observed for the combined selection of “drink slowly during the first 30 min” and “put the drink down between sips” (coefficient-4.46 g; 95% CI - 8.90 to-0.02; p = 0.049). However, the combined selection was not clearly different from selecting “drink slowly during the first 30 min” alone (coefficient 0.16 g; 95% CI-6.65 to 6.98; p = 0.961), and these patterns were not clearly reflected in 120-min total intake (Supplementary Table S6). In the complementary four-category model, the overall association of strategy pattern with 0–30 min intake was suggestive but not definitive (overall p = 0.075), reinforcing the exploratory nature of these findings. These analyses were exploratory and non-causal because strategy choices were self-selected and overlapping within a package intervention.

Usual drinking and alcohol intake in prior 7 days are shown as mean ± standard deviation (SD) and median [first quartile (Q1), third quartile (Q3)]. BMI, body mass index; ADH1B, alcohol dehydrogenase 1B; ALDH2, aldehyde dehydrogenase 2. Genotype distributions are shown because the observed ALDH2 imbalance informed interpretation of genotype-adjusted sensitivity analyses; Fisher’s exact test results are reported only in the Results text and were not used for baseline variable selection.

**Figure 1.**
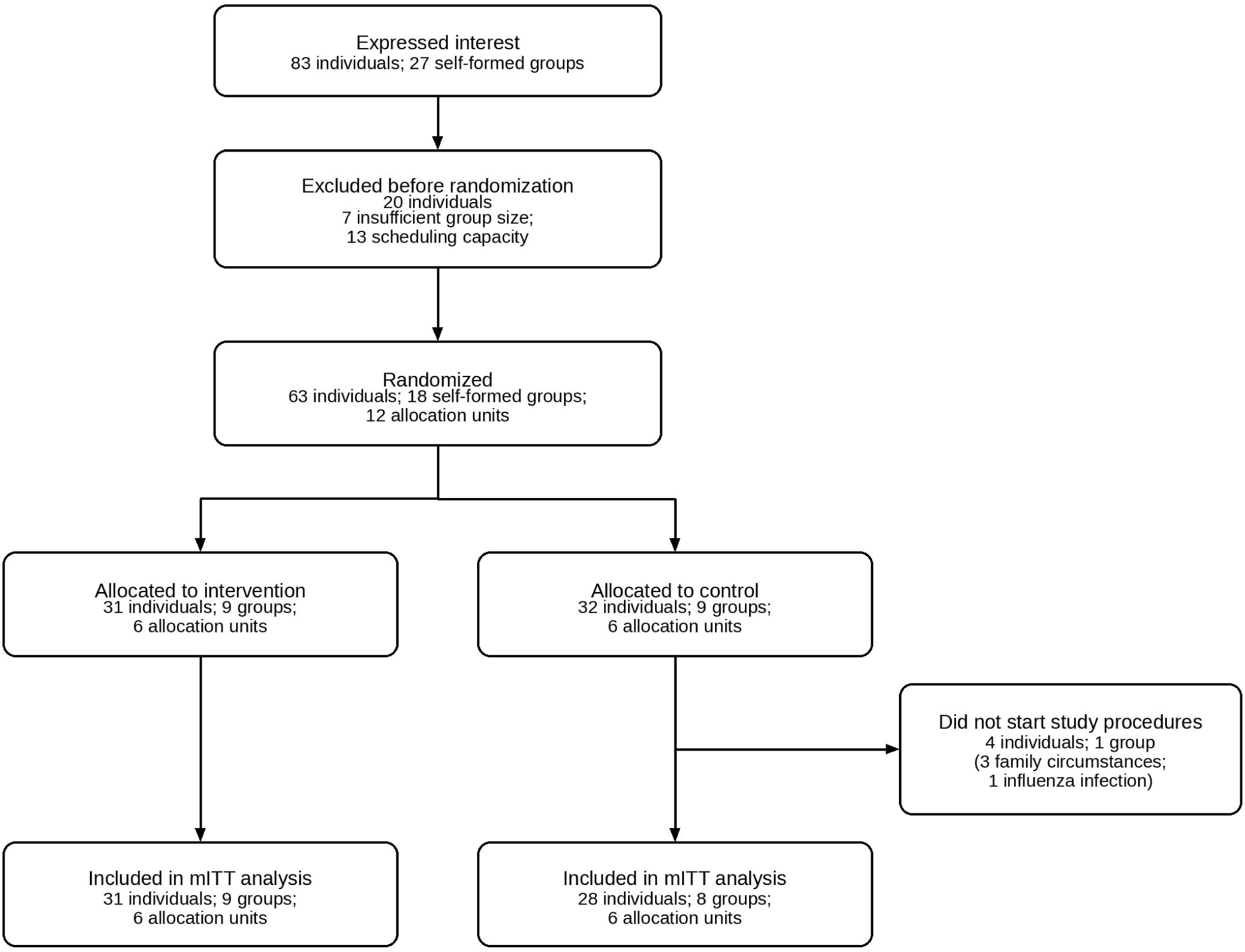
Flow diagram. The 20 individuals excluded before randomization belonged to 9 self-formed groups. One withdrawn control self-formed group was nested within an allocation unit that also contained another participating self-formed group; therefore, all 12 allocation units remained analyzable.

The four participants who withdrew before study procedures had not yet been informed of their assigned condition.

**Figure 2.**
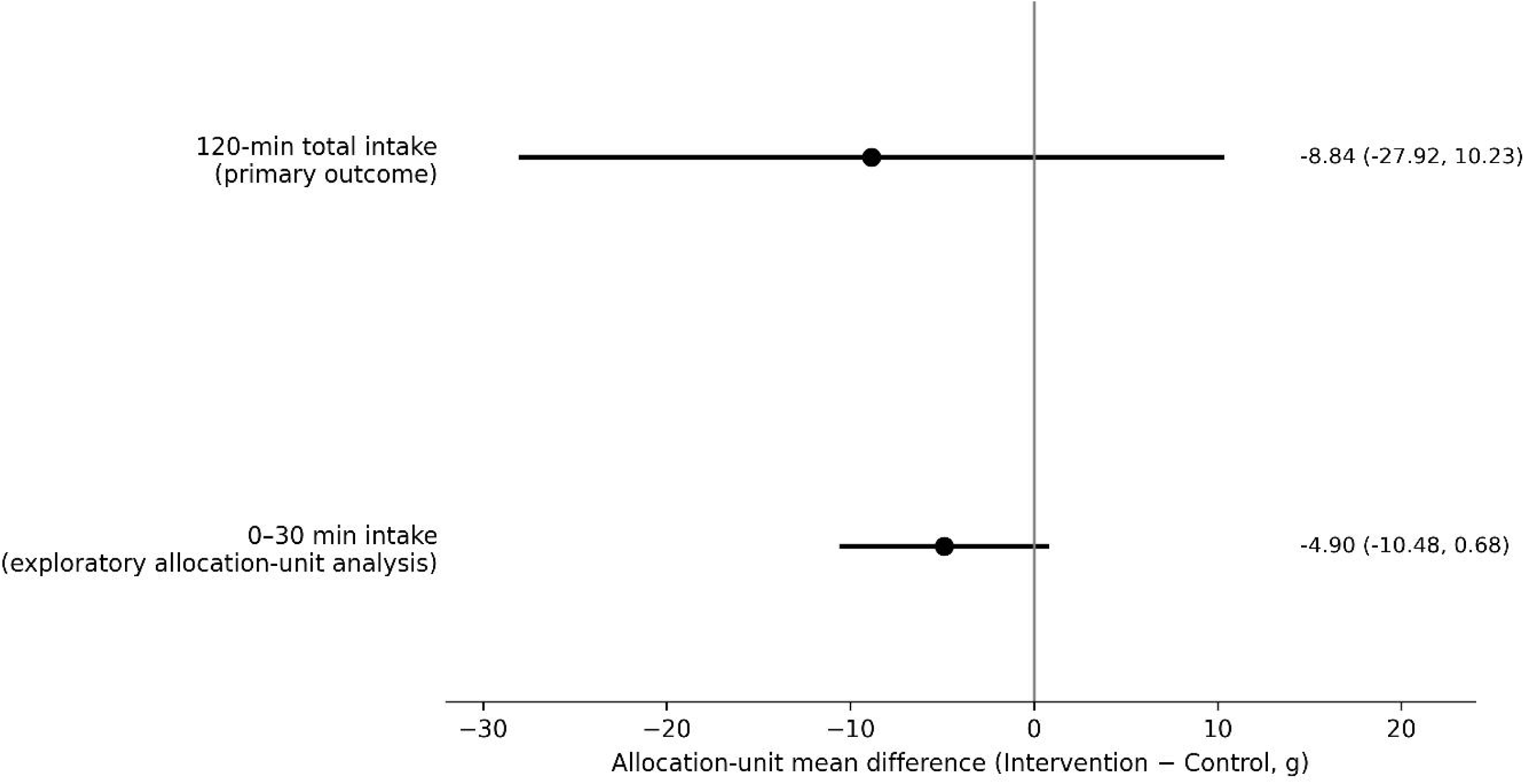
Allocation-unit estimates for the primary outcome and exploratory 0–30 min intake. Points show allocation-unit mean differences (intervention – control), and horizontal bars show t-based 95% confidence intervals for paired block differences; p values are reported in the text and Supplementary Table S2.

## 4. Discussion

This pilot cluster randomized trial showed that session delivery, beverage measurement, and next-day follow-up could be completed once sessions began. The main feasibility constraints arose before randomization: participants had to enroll as self-formed social groups and those groups had to be fitted into limited session slots. In allocation-unit analyses aligned with the unit of randomization, intervention–control point estimates were below zero for both 120-min total alcohol intake and the exploratory 0–30 min interval, but the confidence intervals were wide because the number of allocation units was small. Session satisfaction was high in both arms, and there was no clear between-arm difference.

This study allowed us to test public-facing drinking advice during an actual group drinking episode rather than relying on retrospective self-report or solitary laboratory paradigms [3–5,8–11]. Public guidance in Japan, the United Kingdom, Australia, and Canada commonly recommends pacing, eating while drinking, and alternating alcoholic with non-alcoholic drinks [12–15], but direct experimental evidence from small-group drinking episodes remains limited [8–11]. The present pilot therefore provides a laboratory platform for evaluating widely disseminated risk-reduction advice under socially interactive conditions. Even without precise effect estimation, this pilot suggests that experimentally testing brief, strategy-based advice during a socially interactive drinking episode is operationally feasible and may be relevant to future public-health evaluation of practical drinking guidance. The high satisfaction scores in both arms also support retaining participant experience or acceptability as a prespecified evaluation domain in a future definitive trial.

The results do not identify a single active strategy. Across all self-selected strategies, uptake was heterogeneous, with no uptake for some strategies and near-universal uptake for others. Within that descriptive pattern, the clearest exploratory signal was for early-pacing-related selections, particularly drinking slowly during the first 30 min. A similar signal was observed for the combined selection of drinking slowly during the first 30 min and putting the drink down between sips, but the combined selection was not clearly different from selecting slow initial drinking alone. Because strategy choices were self-selected, overlapping, and not randomized, these findings should be interpreted as hypothesis-generating rather than strategy-specific causal effects. The intervention tested here was a package intervention combining information provision, participant self-selection, and explicit commitment to same-day implementation; accordingly, the strategy-related findings are best understood as within-package exploratory signals rather than evidence for isolated strategy effects. Alcohol-metabolism genotype is another important design consideration in East Asian samples. Despite eligibility criteria intended to enrich for experienced, non-flushing drinkers, substantial ALDH2 imbalance remained between arms. Adjustment for ADH1B and ALDH2 materially changed the intervention effect estimate, and the genotype-adjusted sensitivity model yielded a larger reduction estimate with a confidence interval excluding zero, whereas exploratory interaction analyses did not show a clear genotype-specific pattern. Together, these findings underscore the value of considering ADH1B and ALDH2 in future East Asian trials and suggest that phenotypic screening alone may not eliminate genotype-related heterogeneity [16,17]. This issue may be particularly relevant in small samples, where randomization may not adequately balance genotype distributions.

This study has several limitations. First, the unit of randomization was the time-slot allocation unit, whereas individual-level mixed models used self-formed groups as the analytic clustering variable; although allocation-unit analyses addressed this mismatch, the number of allocation units was small. Second, allocation-unit sizes varied, so the primary allocation-unit analyses estimate an unweighted allocation-unit average effect rather than a participant-average effect [21]; block-size-weighted exact sensitivity analyses are reported in the supplementary materials, but these do not represent an exact participant-average treatment effect when arm sizes differ within blocks. Third, masking could not be maintained throughout the drinking session because the intervention required condition-specific materials and a commitment declaration; however, the primary outcome was based on staff-recorded beverage residual volumes rather than subjective ratings. Fourth, genotype adjustment was not prespecified in the original protocol and should therefore be interpreted as a sensitivity analysis prompted by the observed ALDH2 imbalance. Fifth, post-randomization losses occurred only in the control arm, albeit before study procedures, which may have influenced estimates in this small pilot trial. Sixth, requiring natural social groups of at least three participants and operating within limited session capacity created practical recruitment and scheduling constraints before randomization. Seventh, intervention uptake varied markedly across strategy categories, further limiting strategy-specific inference.

Finally, this was a single-site pilot trial involving experienced drinkers who could participate in social groups and screened as non-flushers; generalizability to solitary drinkers, lighter drinkers, or clinical populations is limited. The unadjusted self-formed-group ICC for 120-min total intake was 0.33, suggesting non-trivial within-group correlation and reinforcing the need to account for clustering explicitly in the design and sample-size planning of a future definitive trial. These limitations mean that the current results are best used to plan a larger trial, including its recruitment approach, analytic strategy, and target precision.

## 5. Conclusions

This pilot showed that brief practical alcohol risk-reduction advice with a commitment element can be delivered feasibly in a socially interactive drinking setting. Primary allocation-unit estimates favored lower alcohol intake but were imprecise. Larger trials are needed to estimate effects more precisely and to evaluate broader public-health applicability, while considering the possible influence of genotype imbalance on effect estimation in East Asian samples.

## 6. List of abbreviations

ADH1B, alcohol dehydrogenase 1B; ALDH2, aldehyde dehydrogenase 2; AUDIT, Alcohol Use Disorders Identification Test; BMI, body mass index; CI, confidence interval; CONSORT, Consolidated Standards of Reporting Trials; ICC, intraclass correlation coefficient; ICD-11, International Classification of Diseases, 11th Revision; mITT, modified intention-to-treat; PBS, protective behavioral strategies; SD, standard deviation; UMIN-CTR, University Hospital Medical Information Network Clinical Trials Registry; VAS, Visual Analog Scale.

## 7. Additional files

Additional file 1: This file contains Supplementary Tables S1–S6 and Appendices A–C, including the registry/protocol outcome mapping, block-level allocation-unit results, genotype-related sensitivity analyses, session-satisfaction analyses, descriptive all-strategy summaries, focused exploratory early-pacing strategy models, intervention-material summaries, beverage and food offerings, safety procedures, and protocol-to-analysis deviations.

Additional file 2: Supplementary Figure S1 shows strategy-selection frequencies within the intervention arm.

Additional file 3: Supplementary Figure S2 reports exploratory interval-specific mixed-model estimates.

Additional file 4: Supplementary Figure S3 shows participant-level mean ± standard deviation (SD) pure alcohol intake by 30-min interval and study arm.

## 8. Declarations

### Ethics approval and consent to participate

The study was approved by the Institutional Ethics Committee for Human Genome and Genetic Research, University of Tsukuba (approval numbers G316 and G316-1). All participants provided written informed consent before participation.

### Consent for publication

Not applicable.

### Availability of data and materials

De-identified participant-level data and analytic code are not publicly posted because the dataset includes small social-group sessions and genotype information. Data and code are available from the corresponding author on reasonable request, subject to ethics approval, applicable institutional requirements, and a data-sharing agreement.

### Authors’ contributions

HY conceived and led the study, including study design, study conduct, project administration, and supervision, and drafted the manuscript. TH contributed to study design, performed the formal analyses, and contributed to validation of the analytic approach. KS contributed to study design and study implementation. MG provided methodological oversight and validation of the statistical aspects of the study. YKu, TK, TF, YKo, KU, and MI contributed to study design, data curation, on-site investigation, project administration, and resources. All authors interpreted the data, critically revised the manuscript for important intellectual content, and approved the final manuscript.

## Supporting information

CONSORT

flow

results

## Acknowledgements

We thank Tetsuya Kodaira (Department of Anesthesiology, University of Tsukuba Hospital), Yuto Iioka, Kentaro Minami, and Takahisa Inaba (Department of Cardiology, University of Tsukuba Hospital), and Yoko Hara and Kaoru Date (University of Tsukuba) for their support with study-day procedures and on-site operations. We also thank the Center for Innovative Medicine and Engineering (CIME), University of Tsukuba Hospital, for institutional support and permission to conduct the study.

## Authors’ information

Not applicable.

