## supplementary for "Practical alcohol risk-reduction advice plus a brief commitment declaration in a social drinking laboratory: a pilot cluster randomized trial"

This supplementary document accompanies the manuscript and provides additional detail on registered outcomes, block-level allocation-unit results, genotype-related sensitivity analyses, session satisfaction, descriptive summaries across all self-selected strategies, focused exploratory within-intervention-arm strategy models, intervention materials, beverage and food offerings, safety procedures, and protocol-to-analysis deviations.

Supplementary Table S1. Outcome mapping from registry/protocol to the current manuscript

| Registered/protocol item | Classification | Reported in this manuscript? | Location / note |
| --- | --- | --- | --- |
| Pure alcohol intake (registry primary outcome) | Primary | Yes | Operationalized here as 120-min total pure alcohol intake in the main manuscript. |
| 30-min interval pure alcohol intake | Derived exploratory outcome | Yes | Main manuscript (exploratory interval analyses). |
| Breath alcohol concentration | Secondary | No | Registry secondary outcome; planned separate report. |
| Blood pressure / heart rate / body temperature | Secondary | No | Registry secondary outcomes; planned separate report. |
| Other registry psychometric outcomes | Secondary | No | Registry secondary outcomes; planned separate report. |
| VAS (including session satisfaction) | Secondary | Partially | Session satisfaction VAS is reported in the main manuscript and Supplementary Table S5; other VAS items are planned for a separate report. |
| Alcohol dehydrogenase 1B / aldehyde dehydrogenase 2 genotypes | Ancillary baseline biomarker | Yes | Not a registered efficacy outcome; reported here as sensitivity-analysis covariates. |

Registry wording was checked against the University Hospital Medical Information Network Clinical Trials Registry (UMIN-CTR) entry. The registry lists the primary outcome broadly as “pure alcohol intake”; in this manuscript, that outcome is operationalized as 120-min total pure alcohol intake, and 0–30 min intake is reported as an exploratory derived outcome. Session satisfaction Visual Analog Scale (VAS), included within the registry secondary VAS outcomes, is reported here as a prespecified secondary participant-experience outcome. No formal progression criteria were prespecified in the protocol.

Abbreviations used in Supplementary Table S1 include BAES, Biphasic Alcohol Effects Scale; SHAS, Subjective High Assessment Scale; PANAS, Positive and Negative Affect Schedule; and AHSS, Alcohol Hangover Severity Scale.

Supplementary Table S2. Block-level allocation-unit means, allocation-unit composition, and block-size-weighted sensitivity analyses

| Outcome | Block | Intervention unit mean (n participants, groups) | Control unit mean (n participants, groups) | Difference (intervention−control, g) | Total participants |
| --- | --- | --- | --- | --- | --- |
| 120-min total intake | 1 | 47.94 (n=5, groups=1) | 76.76 (n=7, groups=2) | -28.82 | 12 |
| 120-min total intake | 2 | 71.67 (n=3, groups=1) | 62.58 (n=8, groups=2) | 9.09 | 11 |
| 120-min total intake | 3 | 44.65 (n=4, groups=1) | 66.47 (n=3, groups=1) | -21.82 | 7 |
| 120-min total intake | 4 | 70.04 (n=7, groups=2) | 72.93 (n=3, groups=1) | -2.89 | 10 |
| 120-min total intake | 5 | 55.25 (n=6, groups=2) | 41.03 (n=3, groups=1) | 14.22 | 9 |
| 120-min total intake | 6 | 49.87 (n=6, groups=2) | 72.70 (n=4, groups=1) | -22.83 | 10 |
| 0–30 min intake | 1 | 16.14 (n=5, groups=1) | 23.47 (n=7, groups=2) | -7.33 | 12 |
| 0–30 min intake | 2 | 18.83 (n=3, groups=1) | 16.28 (n=8, groups=2) | 2.56 | 11 |
| 0–30 min intake | 3 | 19.70 (n=4, groups=1) | 22.60 (n=3, groups=1) | -2.90 | 7 |
| 0–30 min intake | 4 | 20.20 (n=7, groups=2) | 29.43 (n=3, groups=1) | -9.23 | 10 |
| 0–30 min intake | 5 | 17.62 (n=6, groups=2) | 18.70 (n=3, groups=1) | -1.08 | 9 |
| 0–30 min intake | 6 | 14.62 (n=6, groups=2) | 26.03 (n=4, groups=1) | -11.41 | 10 |

Supplementary Table S2 lists allocation-unit composition and block-level means. For 120-min total intake, the unweighted block-paired mean difference was -8.84 g and the block-size-weighted summary difference was -8.95 g; exact sign-flip p values were 0.281 for both analyses. For exploratory 0–30 min intake, the corresponding values were -4.90 g and -5.02 g, with exact sign-flip p values of 0.094 and 0.125. The block-size-weighted summary difference weights each block-specific intervention–control difference by the total number of analyzed participants in the block, where D_b denotes the block-specific intervention–control difference and N_b denotes the total number of analyzed participants in block b; the corresponding test statistic is T = Σ(N_b × D_b) over the 2^6 possible sign permutations. This sensitivity analysis gives larger blocks more influence but is not an exact participant-average treatment effect when arm sizes differ within blocks. All 12 allocation units remained analyzable because the withdrawn control self-formed group was nested within an allocation unit that also contained another participating group.

Supplementary Table S3. Exploratory genotype interaction analyses from Model 4-based mixed-effects models with random intercepts for self-formed groups (coefficient-level Wald p values and omnibus likelihood-ratio tests)

| Model / term | Estimate | 95% confidence interval | Coefficient-level Wald p | Omnibus likelihood-ratio test p / note |
| --- | --- | --- | --- | --- |
| Intervention × ALDH2*1/*2 | -1.54 | -37.21 to 34.14 | 0.933 | Omnibus 1-df LRT p = 0.933. |
| Intervention × ADH1B*1/*2 | 25.01 | -25.44 to 75.47 | 0.331 | Omnibus 2-df LRT p = 0.336. |
| Intervention × ADH1B*2/*2 | 34.71 | -14.09 to 83.50 | 0.163 | Omnibus 2-df LRT p = 0.336. |
| Self-formed-group intraclass correlation coefficient (ICC) for 120-min total intake (unadjusted mixed model) | 0.33 | — | — | Intraclass correlation coefficient from the unadjusted participant-level mixed model. |

Estimate values are interaction coefficients for the intervention-by-genotype terms, relative to the reference genotype categories (alcohol dehydrogenase 1B [ADH1B] *1/*1 and aldehyde dehydrogenase 2 [ALDH2] *1/*1). ADH1B is represented by two indicator terms and is therefore accompanied by a 2-degree-of-freedom omnibus likelihood-ratio test; ALDH2 is represented by one indicator term and is therefore accompanied by a 1-degree-of-freedom omnibus likelihood-ratio test.

Supplementary Table S4. Descriptive summaries of self-selected strategy uptake within the intervention arm

| Strategy | Selected n (%) | 0–30 min crude difference (selected − not selected, g) | 120-min total crude difference (selected − not selected, g) | Descriptive note |
| --- | --- | --- | --- | --- |
| Decide today's drinking amount in advance (optional pure-alcohol target entry) | 0/31 (0.0%) | — | — | No uptake observed. |
| Drink slowly during the first 30 min | 16/31 (51.6%) | -6.09 | -9.05 | Moderate uptake observed. |
| Eat while drinking | 29/31 (93.5%) | 8.52 | 11.21 | Near-universal uptake; comparisons with non-selection are highly unstable. |
| Alternate with water/non-alcoholic drinks | 15/31 (48.4%) | 1.54 | 8.40 | Moderate uptake observed. |
| Choose lower-alcohol beverages | 6/31 (19.4%) | -2.98 | -6.33 | Low uptake observed. |
| Put the drink down between sips | 21/31 (67.7%) | 1.89 | -3.32 | Common uptake observed. |

Note. Strategy selections were made within the package intervention, were self-selected, and were not mutually exclusive. Counts and percentages are provided for description only and should not be interpreted as strategy-specific effects.

Supplementary Table S5. Session satisfaction Visual Analog Scale (VAS) analyses

| Analysis | Intervention | Control | Estimate (intervention−control) | 95% confidence interval | p value / sample size |
| --- | --- | --- | --- | --- | --- |
| Participant-level descriptive comparison | 87.8 ± 12.7 | 91.7 ± 10.0 | -3.88 points | -9.81 to 2.05 | 0.196; 59 participants |
| Allocation-unit analysis | — | — | -3.98 points | -18.77 to 10.82 | 0.531*; 6 blocks |
| Participant-level mixed model | — | — | -3.33 points | -11.50 to 4.83 | 0.424; 59 participants |

Session satisfaction VAS was prespecified as a secondary outcome. The block-size-weighted summary difference was -4.88 points, with an exact sign-flip p value of 0.531. Intervention and Control columns are shown only for the descriptive participant-level comparison. Allocation-unit and mixed-model rows report between-arm contrast estimates only.

Supplementary Table S6. Focused exploratory within-intervention-arm models for early-pacing-related selections

| Outcome | Selection / contrast | Adjusted coefficient (g) | 95% confidence interval | p value |
| --- | --- | --- | --- | --- |
| 0–30 min intake | Selected “drink slowly during the first 30 min” | -5.88 | -10.07 to -1.69 | 0.008 |
| 120-min total intake | Selected “drink slowly during the first 30 min” | -8.74 | -24.54 to 7.06 | 0.267 |
| 0–30 min intake | Selected both “drink slowly during the first 30 min” and “put the drink down between sips” | -4.46 | -8.90 to -0.02 | 0.049 |
| 120-min total intake | Selected both “drink slowly during the first 30 min” and “put the drink down between sips” | -7.91 | -23.72 to 7.91 | 0.314 |
| 0–30 min intake | Four-category model: combined selection versus selection of “drink slowly during the first 30 min” only | 0.16 | -6.65 to 6.98 | 0.961 |
| 120-min total intake | Four-category model: combined selection versus selection of “drink slowly during the first 30 min” only | -3.54 | -28.81 to 21.72 | 0.775 |

Models were fitted within the intervention arm and adjusted for usual drinking and alcohol intake during the prior 7 days. The combined-selection indicator denotes participants who selected both “drink slowly during the first 30 min” and “put the drink down between sips.” In the four-category model, category sizes were n = 6 for neither selection, n = 9 for selection of “put the drink down between sips” only, n = 4 for selection of “drink slowly during the first 30 min” only, and n = 12 for the combined selection of both strategies. The overall p value for the strategy factor was 0.075 for 0–30 min intake and 0.548 for 120-min total intake. These analyses were exploratory and non-causal because strategy selections were self-selected and overlapping within a package intervention.

Supplementary Appendix A. Summaries of intervention and control materials

Intervention sessions delivered a package intervention comprising practical pre-drinking strategy information, participant self-selection of one or more same-day strategies, and an explicit commitment declaration. Control sessions delivered alcohol-related knowledge only. Core intervention content asked participants to choose at least one strategy for use during the session: deciding the amount to drink that day in advance (with an optional planned pure-alcohol target entry), drinking slowly during the first 30 min, eating while drinking, alternating alcoholic drinks with water or non-alcoholic beverages, choosing lower-alcohol beverages, and putting the drink down between sips. Condition-specific information sessions lasted approximately 5–10 min. This appendix summarizes the materials and delivery elements rather than reproducing the full source documents.

Additional material-specific details are as follows. Alcoholic options included beer, canned ready-to-drink cocktails, canned sparkling fruit or plum-wine drinks, and red or white wine served in 100-mL pours. Pure alcohol calculations used the label alcohol content of the specific product offered that day and the recorded volume consumed. Alcohol-free options included alcohol-free beer-style and lemon beverages, green tea, water, and carbonated water. Brand names are omitted.

Supplementary Appendix B. Summary of safety monitoring and stopping procedures

Additional operational points not repeated in the main manuscript were a 60-min post-drinking recovery observation period, continued safety checks at 150 and 180 min, restroom accompaniment as needed to reduce fall risk, and taxi return according to predefined departure procedures.

Supplementary Appendix C. Protocol-to-analysis deviations

The original protocol described self-formed groups as clusters and envisaged primary analyses at the group/participant level. Because the operational unit of randomization was the time-slot allocation unit, the present report centers allocation-unit analyses as the primary inferential analyses and treats individual-level mixed-effects models as sensitivity analyses. Genotype adjustment is reported as a biologically motivated sensitivity analysis because substantial ALDH2 imbalance was observed despite phenotypic screening for flushing.
