## Supplementary figures and images for "Practical alcohol risk-reduction advice plus a brief commitment declaration in a social drinking laboratory: a pilot cluster randomized trial"

### figureS1

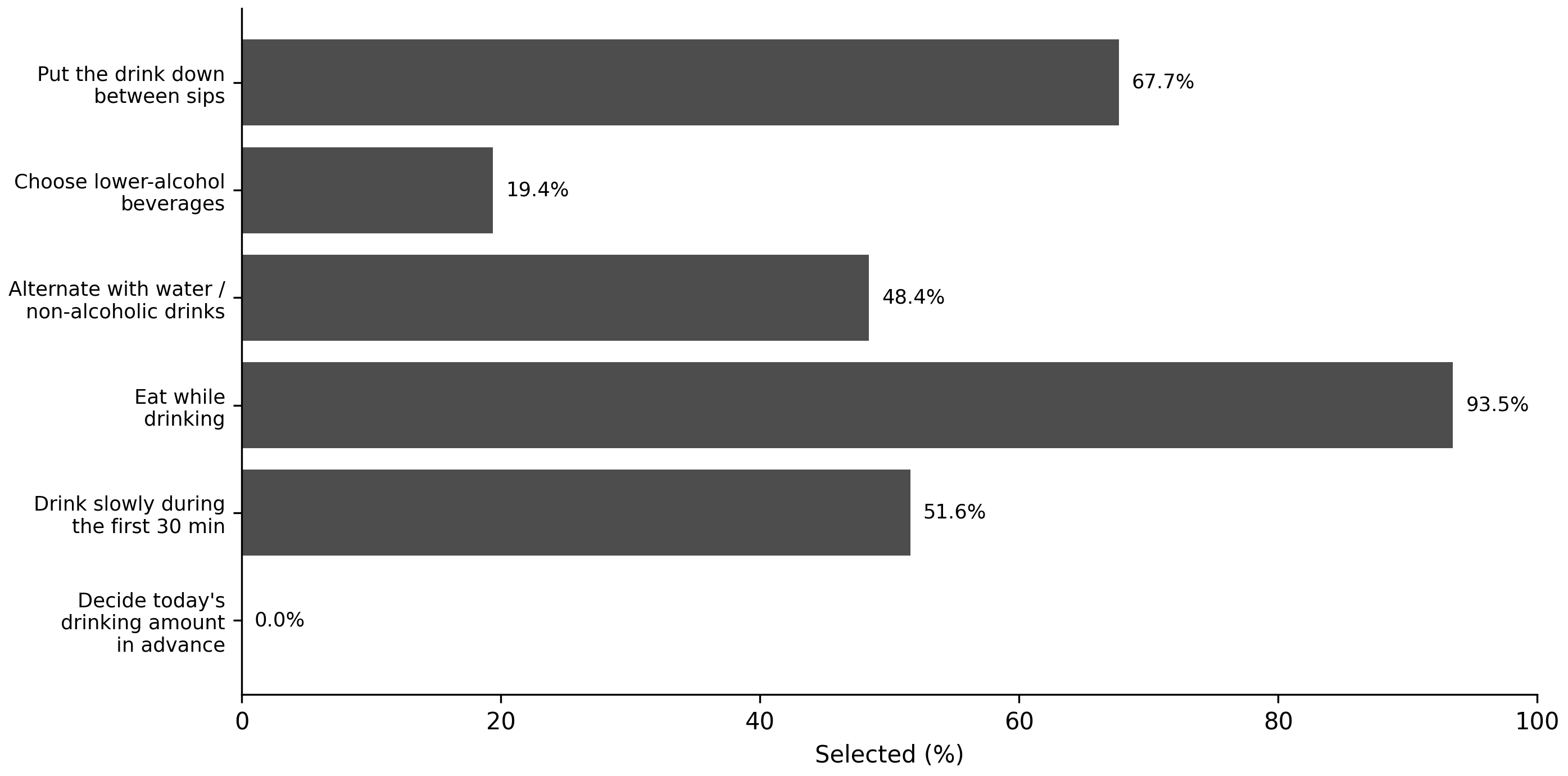

### figureS2

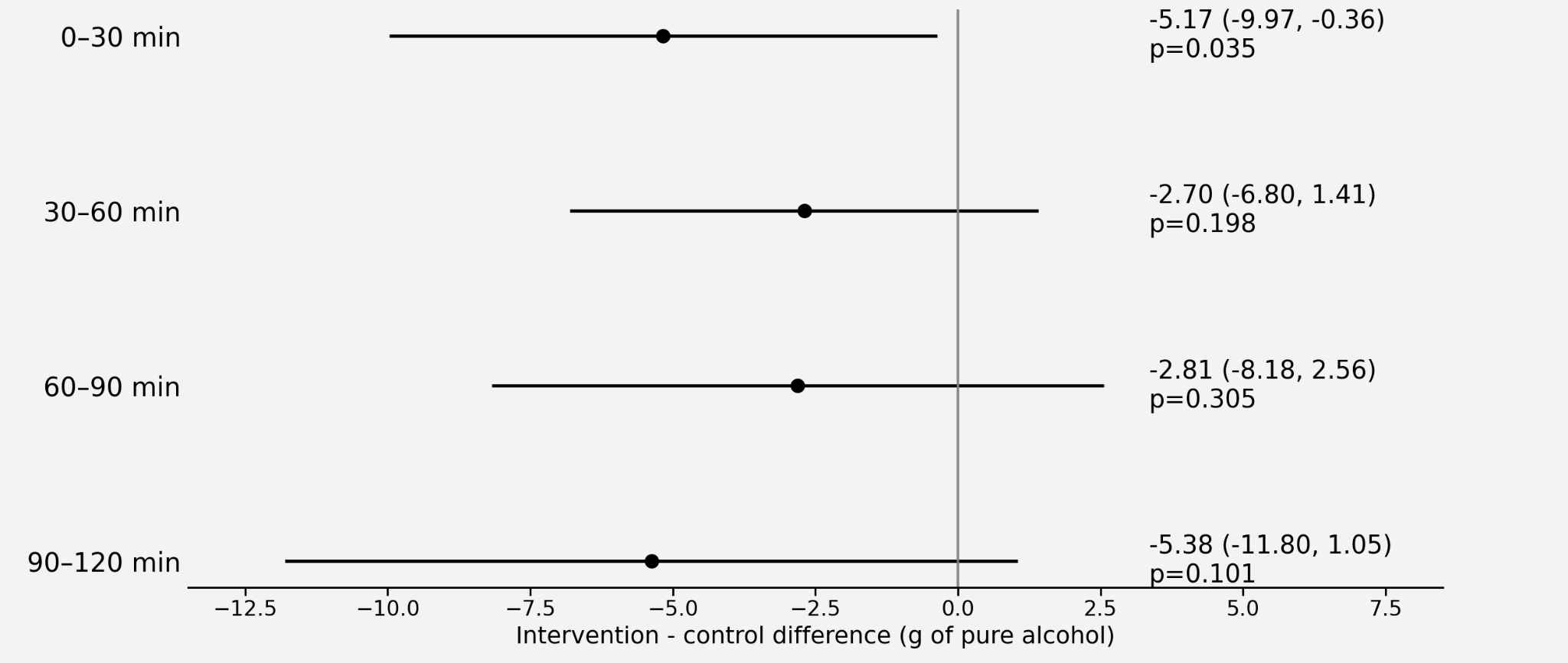

### figureS3

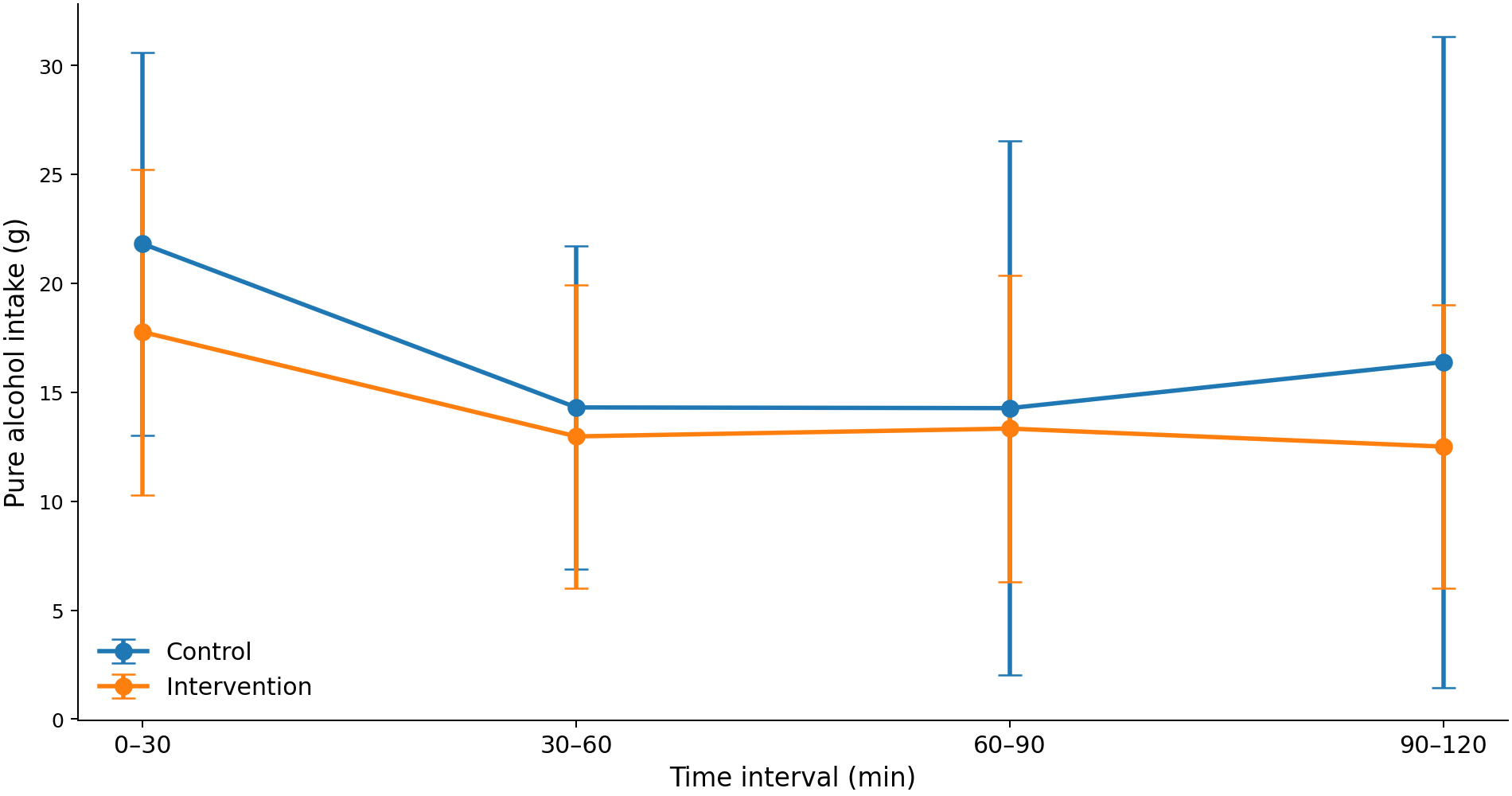

### flow

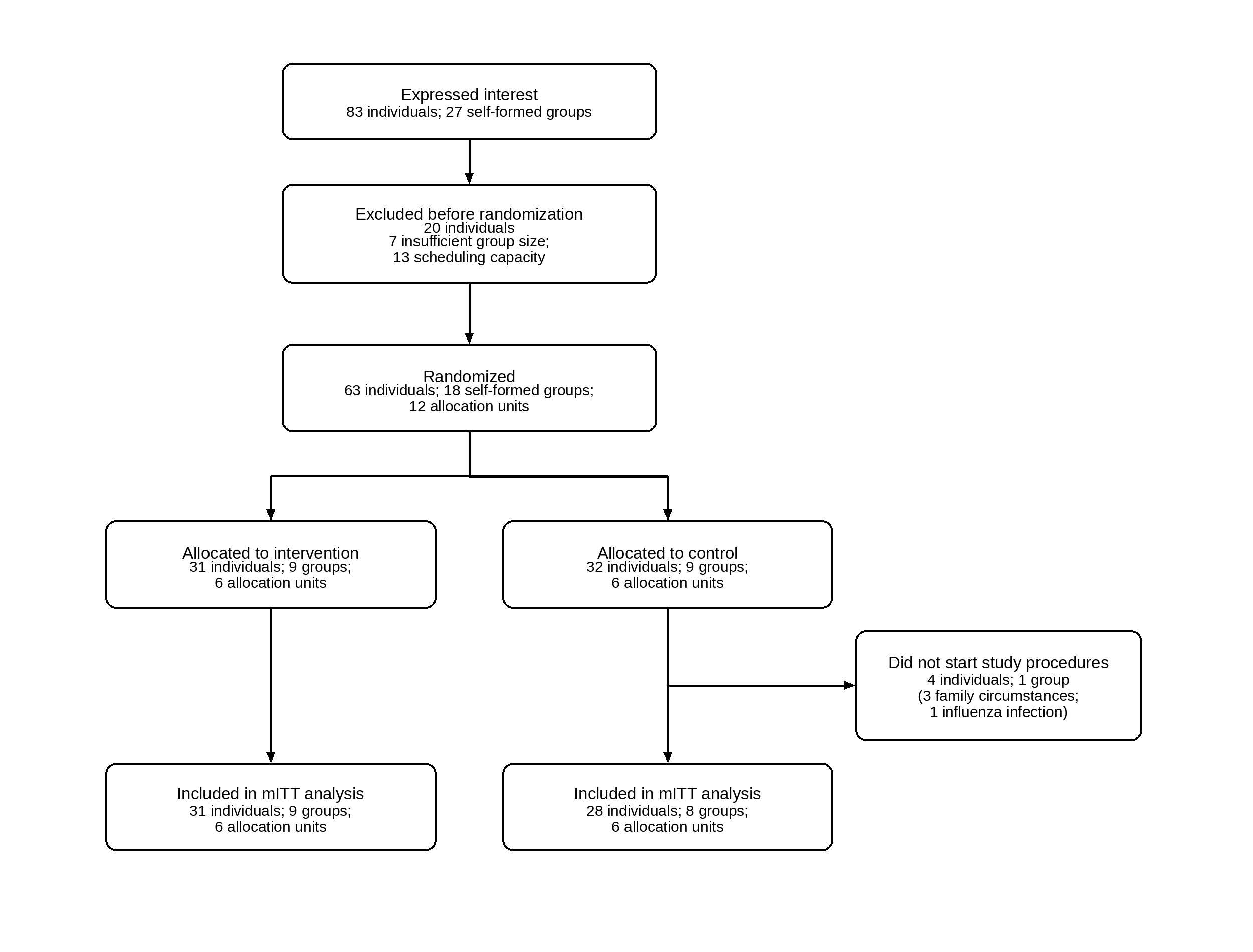

### results

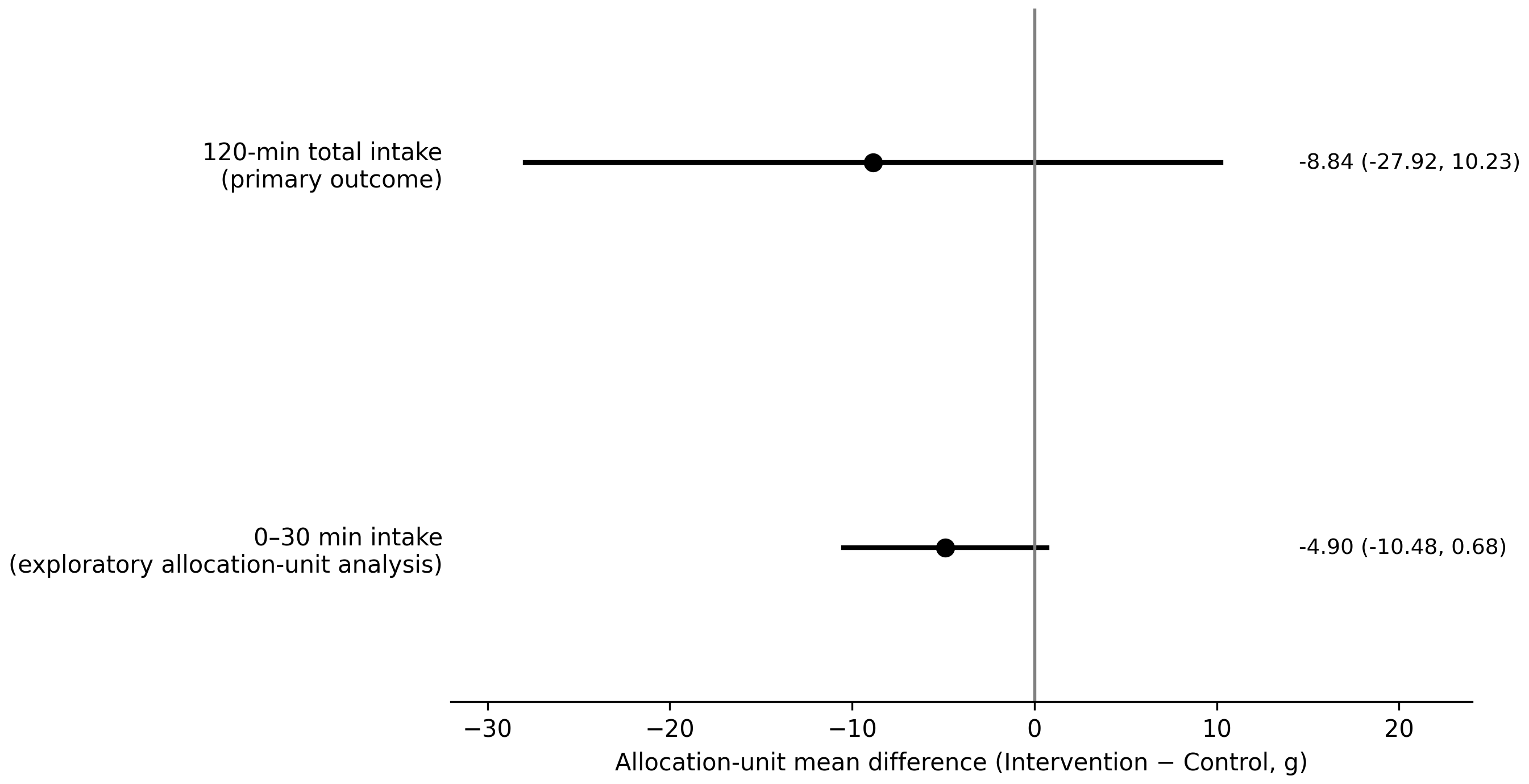
