## Supplementary material for "Practical alcohol risk-reduction advice plus a brief commitment declaration in a social drinking laboratory: a pilot cluster randomized trial": CONSORT

**CONSORT 2010 pilot and feasibility extension checklist**

Filled for the current BMC Public Health submission package

| **Topic** | **Item** | **Checklist item** | **Reported location** |
| --- | --- | --- | --- |
| **Title and abstract** | | | |
| Title | 1a | Identification as a pilot or feasibility randomised trial in the title | Main manuscript, title |
| Abstract | 1b | Structured summary of pilot trial design, methods, results, and conclusions | Main manuscript, Abstract |
| **Introduction** | | | |
| Background and rationale | 2a | Scientific background and explanation of rationale for future definitive trial, and reasons for randomised pilot trial | Main manuscript, Background |
| Objectives | 2b | Specific objectives or research questions for pilot trial | Main manuscript, Background, final paragraph |
| **Methods** | | | |
| Trial design | 3a | Description of pilot trial design (such as parallel, factorial) including allocation ratio | Main manuscript, Methods 2.1 |
| Trial design | 3b | Important changes to methods after pilot trial commencement, with reasons | Additional file 1, Appendix C |
| Participants | 4a | Eligibility criteria for participants | Main manuscript, Methods 2.2 |
| Participants | 4b | Settings and locations where the data were collected | Main manuscript, Methods 2.1 and 2.4 |
| Participants | 4c | How participants were identified and consented | Main manuscript, Methods 2.2; Declarations: Ethics approval and consent to participate |
| Interventions | 5 | The interventions for each group with sufficient details to allow replication, including how and when they were actually administered | Main manuscript, Methods 2.4; Additional file 1, Appendix A |
| Outcomes | 6a | Completely defined prespecified assessments or measurements to address each pilot trial objective specified in 2b, including how and when they were assessed | Main manuscript, Methods 2.5; Results 3.3–3.5; Additional file 1, Tables S1–S6 |
| Outcomes | 6b | Any changes to pilot trial assessments or measurements after the pilot trial commenced, with reasons | Additional file 1, Table S1 and Appendix C |
| Outcomes | 6c | If applicable, prespecified criteria used to judge whether, or how, to proceed with future definitive trial | Main manuscript, Methods 2.6; Additional file 1, Table S1 |
| Sample size | 7a | Rationale for numbers in the pilot trial | Main manuscript, Methods 2.1 |
| Sample size | 7b | When applicable, explanation of any interim analyses and stopping guidelines | Main manuscript, Methods 2.4 (no interim analyses) |
| Randomisation | 8a | Method used to generate the random allocation sequence | Main manuscript, Methods 2.3 |
| Randomisation | 8b | Type of randomisation(s); details of any restriction (such as blocking and block size) | Main manuscript, Methods 2.3 |
| Allocation concealment | 9 | Mechanism used to implement the random allocation sequence and any steps taken to conceal the sequence until interventions were assigned | Main manuscript, Methods 2.3–2.4 |
| Implementation | 10 | Who generated the random allocation sequence, who enrolled participants, and who assigned participants to interventions | Main manuscript, Methods 2.3; Declarations: Authors’ contributions |
| Blinding | 11a | If done, who was blinded after assignment to interventions and how | Main manuscript, Methods 2.4 |
| Blinding | 11b | If relevant, description of the similarity of interventions | Not applicable |
| Statistical methods | 12 | Methods used to address each pilot trial objective, whether qualitative or quantitative | Main manuscript, Methods 2.6; Results 3.3–3.5; Additional file 1, Tables S2–S6 |
| **Results** | | | |
| Participant flow | 13a | For each group, the numbers of participants who were approached and/or assessed for eligibility, randomly assigned, received intended treatment, and were assessed for each objective | Main manuscript, Results 3.1; Figure 1 |
| Participant flow | 13b | For each group, losses and exclusions after randomisation, together with reasons | Main manuscript, Results 3.1; Figure 1 |
| Recruitment | 14a | Dates defining the periods of recruitment and follow-up | Main manuscript, Methods 2.1; Results 3.1 |
| Recruitment | 14b | Why the pilot trial ended or was stopped | Not applicable; the pilot trial ended as planned. |
| Baseline data | 15 | A table showing baseline demographic and clinical characteristics for each group | Main manuscript, Table 1 |
| Numbers analysed | 16 | For each objective, number of participants (denominator) included in each analysis | Main manuscript, Results 3.1 and 3.3–3.5; Tables 1–2; Additional file 1, Tables S2–S6 |
| Outcomes and estimation | 17 | For each objective, results including expressions of uncertainty (such as 95% confidence interval) for any estimates | Main manuscript, Results 3.3–3.5; Table 2; Figure 2; Additional files 1–4 |
| Ancillary analyses | 18 | Results of any other analyses performed that could be used to inform the future definitive trial | Main manuscript, Results 3.4–3.5; Additional files 1–4 |
| Harms | 19 | All important harms or unintended effects in each group | Main manuscript, Methods 2.4; Results 3.1; Discussion (limitations); Additional file 1, Appendix B |
| Other unintended consequences | 19a | If relevant, other important unintended consequences | Not applicable |
| **Discussion** | | | |
| Limitations | 20 | Pilot trial limitations, addressing sources of potential bias and remaining uncertainty about feasibility | Main manuscript, Discussion (limitations) |
| Generalisability | 21 | Generalisability (applicability) of pilot trial methods and findings to future definitive trial and other studies | Main manuscript, Discussion and Conclusions |
| Interpretation | 22 | Interpretation consistent with pilot trial objectives and findings, balancing potential benefits and harms, and considering other relevant evidence | Main manuscript, Discussion |
| Progression | 22a | Implications for progression from pilot to future definitive trial, including any proposed amendments | Main manuscript, Discussion and Conclusions |
| **Other information** | | | |
| Registration | 23 | Registration number for pilot trial and name of trial registry | Main manuscript, Abstract (Trial registration); Methods 2.7 |
| Protocol | 24 | Where the pilot trial protocol can be accessed, if available | Additional file 1, Table S1 and Appendix C |
| Funding | 25 | Sources of funding and other support (such as supply of drugs), role of funders | Main manuscript, Declarations: Funding |
| Ethics approval | 26 | Ethical approval or approval by research review committee, confirmed with reference number | Main manuscript, Declarations: Ethics approval and consent to participate |

*Source used for item wording and structure: Eldridge SM et al. CONSORT 2010 statement: extension to randomised pilot and feasibility trials. BMJ. 2016;355:i5239.*
